# Barriers in accessing healthcare during the COVID-19 pandemic: analysis of the Virus Watch community cohort study

**DOI:** 10.1101/2024.02.15.24302762

**Authors:** Dee Menezes, Chloë Siegele-Brown, Alexei Yavlinsky, Vincent Nguyen, Sarah Beale, Isobel Braithwaite, Rachel Burns, Sam Tweed, Youssof Oskrochi, Thomas Byrne, Jana Kovar, Wing Lam Erica Fong, Andrew Hayward, Robert W Aldridge, the Virus Watch study

## Abstract

**Background:** Differential barriers to accessing healthcare contribute to inequitable health outcomes. This study aims to describe the characteristics of individuals who experienced barriers, and what those barriers were, during the COVID-19 pandemic.

**Methods:** We analysed data from Virus Watch: an online survey-based community study of households in England and Wales. The primary outcome was reported difficulty accessing healthcare in the previous year.

**Results:** Minority ethnic participants reported difficulty accessing healthcare more than White British participants (41.6% vs 37%), while for migrants this was at broadly similar levels to non-migrants. Those living in the most deprived areas reported difficulty more than those living in the least deprived quintile (45.5% vs. 35.5%). The most frequently reported barrier was cancellation/disruption of services due to the COVID-19 pandemic (72.0%) followed by problems with digital or telephone access (21.8%). Ethnic minority participants, migrants, and those from deprived areas more commonly described “insufficient flexibility of appointments” and “not enough time to explain complex needs” as barriers.

**Conclusions:** Minority ethnic individuals and those living in deprived areas were more likely to experience barriers to healthcare during the COVID-19 pandemic, and it is essential they are addressed as services seek to manage backlogs of care.

## Introduction

Inequities in health outcomes related to COVID-19 have been evident since the first few months of the pandemic. Higher rates of COVID-19 disease and death were seen in minoritised groups such as ethnic minorities, migrants and those living in areas of higher deprivation, when compared with those of white ethnicity, non-migrants and those living in areas with less deprivation^1^. These characteristics can overlap and compound health inequities at intersections^2^ and often relate to phenomena beyond the individual such as structural factors including occupational risks^3,4^ and household overcrowding^5^. Health systems can act to both redress health inequities, but also inadvertently reinforce them, leading to differential access to healthcare occurring across demographic groups.

Before the pandemic, there was evidence of inequities in access to healthcare experienced by minoritised groups ^6–10^ and further evidence since suggests that this gap widened during the pandemic ^11–13^. For example, one study showed that existing pre-pandemic differences in consultation rates between migrants and non-migrants grew by a further 11% during the first year of the pandemic in England ^11^.

While some research has examined difficulties in accessing healthcare during the COVID-19 pandemic, much has focused on a single characteristic, rather than taking into account the multitude of ways disadvantage can affect healthcare access. Furthermore, to our knowledge, no literature to date has examined the specific barriers experienced by minoritised groups during the COVID-19 pandemic.

To describe barriers in access to healthcare, we adapted the Médecins du Monde/Doctors of the World (DOTW) healthcare access survey tool to answer two research questions in the UCL Virus Watch cohort study: 1) were people from minority ethnic backgrounds, migrants, and people living in more deprived areas more likely to report difficulties in accessing healthcare between January 2021 and January 2022, compared to White British people, non-migrants, and less deprived groups, respectively? 2) During the same period were there differences in the nature of the barriers faced by these groups when accessing healthcare, with respect to the same comparator groups?

## Methods

Ethics Approval and Consent Virus Watch was approved by the Hampstead NHS Health Research Authority Ethics Committee: 20/HRA/2320, and conformed to the ethical standards set out in the Declaration of Helsinki. All participants provided informed consent for all aspects of the study.

Virus Watch is a prospective household community cohort study in England and Wales that recruited participants from June 2020 to March 2022. The full protocol for the study has been described previously^14^. Briefly, the study recruited participants through a campaign using general post, leaflets, social media, letters and SMS from General Practices all directing potential participants to the study website for information and consent. The study eligibility criteria were: ordinarily residing in England or Wales, living in a household between one and six people (the study was restricted to a maximum of six household members due to limitations on survey infrastructure), internet and email access, and at least one household member being able to read and complete surveys in English.

Participants self-reported their demographics in a baseline survey, including their ethnicity according to 2011 Office of National Statistics (ONS) Census categories during study enrolment, and responses were subsequently aggregated into one of: Black, Mixed, Other Ethnicity, Other Asian, South Asian, White British, White Irish, or White Other. For the purposes of this analysis, participants were considered to be of minority ethnic background if they were not in the White British category. Participants also self-reported their country of birth which was operationalised as a binary variable (UK born/Not UK Born). Socioeconomic deprivation was approximated using local area (Lower Super Output Area) Index of Multiple Deprivation (IMD) quintiles corresponding to the postcodes provided by participants at enrolment, where quintile 1 corresponds to the most deprived 20% of areas and quintile 5 to the least deprived 20%.

After enrolment, participants were emailed a weekly survey in which they reported acute infection symptoms and COVID-19 testing and vaccinations plus monthly surveys, the theme of which was determined ad hoc by the study team and based upon timely policy and participant relevant research questions. In the January 2022 survey, participants were asked questions related to healthcare access. We refer to this survey in the rest of this analysis as the “Access survey”.

Virus Watch participants who answered the January 2022 Access survey had to be aged 18 or over at enrolment. The primary outcome, “Difficulty accessing healthcare”, was assessed by a “Yes” response to the question: “*During the last 12 months, have you experienced any obstacles/barriers when accessing healthcare services?*”. Other possible responses to this question included: ”No – I have had good/easy access to healthcare services”, ”No – I have not tried to access healthcare services”, and “NA/Prefer not to say”. We considered an individual to have tried to access healthcare if they responded either “Yes” or ”No – I have had good/easy access to healthcare services”. Participants answering “Yes” were then able to select from 18 possible reasons for encountering barriers, choosing as many options desired. The list of barriers was adapted from the DOTW service user questionnaire, with additional options added to capture changes in healthcare access during the pandemic (Box 1).

We described the entire Virus Watch Cohort as of January 2022 and the Access survey participants in terms of age group, sex, minority ethnicity, migration status, and IMD quintile, adding the corresponding figures from the 2021 England and Wales Census for comparison to the general population. We then calculated the proportion of respondents who reported the primary outcome, and examined the distribution of reasons given for difficulty accessing healthcare, calculating the proportion selecting each reason by minority ethnicity, migration status, and IMD quintile, along with corresponding 95% confidence intervals. Lastly, these characteristics were also used in a series of regression models to test their association with the outcome. We included a compound term of minority ethnicity and migration status, as we expected that a change in one of these factors would impact the strength of association in the others.

#### Box 1

**List of reasons for difficulty accessing healthcare presented in survey question**

- Services disrupted or cancelled due to COVID-19 pandemic
- Difficult to attend appointments at the time they are offered, e.g. due to work or childcare
- Not enough flexibility of appointments
- Too far away from where I live
- Too many different services/locations to attend
- Fear of not being listened to
- Previous mistreatment
- Previous experience of discrimination or stigma
- Not enough time to explain complex needs
- Problems with digital and/or phone access
- Not knowing the healthcare system
- Difficult understanding the services offered
- Problems with documentation and administration
- Problems with language or literacy
- Worries about being charged
- Fear of immigration enforcement/arrest
- Denied access
- Other (please describe below)

## Results

### Sample characteristics

There were 50,183 participants who were aged 18 years or older in the Virus Watch cohort by 31st January 2022. Of this, 15,568 responded to the Access survey. Respondents tended to be both older and have a larger proportion of White British individuals, when compared with the general population in the 2021 Census and the Virus Watch cohort (Table 1). There was a larger proportion of Access survey respondents who were living in less deprived IMD quintiles. For example, 32% of Access survey respondents lived in the least deprived IMD quintile compared with 26 and 19.7% in the Virus Watch cohort and in the 2021 census, respectively.

**Table 1:** Baseline characteristics of all Virus Watch participants and those who responded to the access survey.

| **Characteristic** | **All Adult**<br>**VirusWatch**<br>**Participants**<br>**(31-Jan-2022)** | **All access survey**<br>**participants** | **ONS**<br>**N= 59,597,542**<br>**(%)** |
| --- | --- | --- | --- |
| **All** | **50,183** | 15,568 | 100 |
| **Age\*** |  |  |  |
| 18-24 | 2,473 (4.9) | 225 (1.4) | 10.6 |
| 25-44 | 11,713 (23.4) | 1,037 (6.7) | 33.1 |
| 45-64 | 19,630 (39.1) | 6,094 (39) | 32.6 |
| 65+ | 16,366 (32.6) | 8,212 (53) | 23.7 |
| **Minority**<br>**ethnicity** |  |  |  |
| White British | 36,116 (72) | 14,218 (91) | 74.4 |
| Minority ethnic | 7,018 (14) | 1,284 (8.2) | 25.7 |
| Missing** | 7,049 (14) | 66 (0.4) | - |
| **Sex** |  |  |  |
| Female | 27,478 (47) | 8,946 (57) | 51.0 |
| Male | 21,666 (37) | 6,622 (43) | 49.0 |
| Missing | 9,312 (16) | 0 | 0 |
| **IMD Quintile** |  |  |  |
| 1 (most<br>deprived) | 5, 042 (10) | 1,139 (7.3) | 18.1 |
| 2 | 7,873 (16) | 2,179 (12) | 20.5 |
| 3 | 9,389 (19) | 3,075 (20) | 20.7 |
| 4 | 11,329 (23) | 4,051 (26) | 20.2 |
| 5 (least<br>deprived) | 12,874 (26) | 4,937 (32) | 19.7 |
| Missing | 3,676 (7.3) | 187 (1.2) | 0 |
| **Migration**<br>**Status** |  |  | (%) |
| UK born | 29,037 (58) | 10,224 (66) | 83.2 |
| Not UK born | 5,052 (10) | 924 (5.9) | 16.8 |
| Missing | 16,094 (32) | 4,420 (28) |  |
| **Region** |  |  | (%) |
| East Midlands | 4,936 (8.4) | 1,487 (9.6) | 8 |
| East of England | 10,535 (18) | 3,158 (20) | 11 |
| London | 9,079 (16) | 1,536 (9.9) | 15 |
| North East | 2,528 (4.3) | 742 (4.8) | 4 |
| North West | 5,562 (9.5) | 1,711 (11) | 12 |
| South East | 9,635 (17) | 3,057 (20) | 16 |
| South West | 3,952 (6.7) | 1,384 (8.9) | 10 |
| Wales | 1,528 (4.3) | 469 (3.0) | 5 |
| West Midlands | 3,014 (5.1) | 925 (5.9) | 10 |
| Yorkshire and The Humber | 3,033 (5.2) | 912 (5.9) | 9 |
| Missing | 42 (<0.1) | 2 (<0.1) | - |

\*Age for a single record was missing. \*\*"Prefer not to say", included in "Missing" accounted for 192 (0.3%) participants.
ONS statistics were sourced as follows:
Overall population and sex:
https://www.ons.gov.uk/peoplepopulationandcommunity/populationandmigration/populationestimates/bulletins/populationandhouseholdestimatesenglandandwales/census2021unroundeddata
Minority ethnicity:
https://www.ons.gov.uk/peoplepopulationandcommunity/culturalidentity/ethnicity/bulletins/ethnicgroupenglandandwales/census2021
Migration:
https://www.ons.gov.uk/peoplepopulationandcommunity/populationandmigration/internationalmigration/bulletins/internationalmigrationenglandandwales/census2021#:~:text=It%20is%20often%20the%20preferred,were%20born%20outside%20the%20UK.
Populations in regions:
https://www.ons.gov.uk/peoplepopulationandcommunity/populationandmigration/populationestimates/bulletins/populationandhouseholdestimatesenglandandwales/census2021

Of the Access survey respondents, 12,237 (79%) reported having tried to access healthcare, while 2,987 (19%) had not tried to access healthcare and 344 (2%) preferred not to say. The following analysis concerns only those participants who reported having tried to access healthcare, the characteristics of whom are shown in Table 2.

**Table 2:** Characteristics of participants who reported having tried to access healthcare.

|  | Participants who reported having tried to access healthcare (N=12,237) | Participants facing a barrier to healthcare access in the previous 12 months |  |
| --- | --- | --- | --- |
| **Sex** |  | **Yes n=4,571** | **No n=7,666** |
| Female | 7102 (58.0) | 2892 (40.7) | 4210 (59.3) |
| Male | 5135 (42.0) | 1679 (32.7) | 3456 (67.3) |
| **Age** |  |  |  |
| 18-24 | 142 (1.2) | 46 (32.4) | 96 (67.6) |
| 25-44 | 734 (6.0) | 336 (45.8) | 398 (54.2) |
| 45-64 | 4705 (38.4) | 1939 (41.2) | 2766 (58.8) |
| 65+ | 6,656 (54.5) | 2250 (33.8) | 4406 (66.2) |
| **Migration status** |  |  |  |
| Not UK born | 699 (5.7) | 275 (39.3) | 424 (60.7) |
| UK born | 8028 (65.6) | 2953 (36.8) | 5075 (63.2) |
| Missing | 3517 (28.7) | 1343 (38.3) | 2167 (61.7) |
| **IMD quintile** |  |  |  |
| 1 (most deprived) | 908 (7.4) | 413 (45.5) | 495 (54.5) |
| 2 | 1703 (13.9) | 671 (39.4) | 1032 (60.6) |
| 3 | 2429 (19.9) | 882 (36.3) | 1547 (63.7) |
| 4 | 3176 (26.0) | 1175 (37) | 2001 (63) |
| 5 (least deprived) | 3873 (31.7) | 1374 (35.5) | 2499 (64.5) |
| Missing | 148 (1.2) | 56 (37.8) | 92 (62.2) |
| **Ethnicity** |  |  |  |
| Minority ethnic | 983 (8.0) | 409 (41.6) | 574 (58.4) |
| White British | 11,208 (91.6) | 4154 (37) | 7054 (62.9) |
| Missing | 46 (0.4) | 8 (17) | 38 (80.9) |

\*344 Prefer not to say, 2,987 did not try to access

### Barriers to healthcare access

Of the 12,237 participants who reported at least one attempt to access healthcare services, 4,571 (37.4%) reported difficulties accessing healthcare (Table 2). Notably, comparatively more participants from minority ethnic backgrounds reported difficulties accessing healthcare than White British participants (41.6% [95% CI 38.6 - 44.7] vs. 37.1% [36.2 - 38.0], and broadly similar proportions of migrants and those born in the UK (39.3% [35.8 - 43.0] vs. 36.8% [35.7 - 37.8], respectively). Participants from the most deprived quintile reported difficulties accessing healthcare more frequently than the least deprived (45.5% [42.3 - 48.7] in the 1st, most deprived IMD quintile vs. 35.5% [34.0 to 37.0] in the 5th). While not the primary focus of this analysis, we also found that higher proportions of 25 - 64 year olds reported difficulties accessing healthcare compared with those in the 65+ age group, and women reported difficulties more frequently than men. We found weak evidence for an association between minority ethnicity (OR 1.1 [1.0-1.3]) and migration status (OR 1.0 [0.9 -1.2]) and reported difficulties accessing healthcare in age and sex-adjusted regression models (Table 3). When including a ethnicity-migration compound variable we found that those who were of minority ethnicity and UK born (“minority ethnicity-UK born”) had a stronger and statistically significant association with the outcome (OR 1.4 (1.1-1.9)). This finding persisted in our sensitivity analysis where IMD quintiles were added to the model (Appendix 3: OR 1.5 (1.1-1.9)).

**Table 3:** Regression models. In this regression model, the outcome is a participant selecting “Yes” to the question “During the last 12 months, have you experienced any obstacles/barriers when accessing healthcare services?” We have included sex, age group, ethnicity in one model, then sex, age group and migration status in the second. In the third model (far right), we have included sex, age group and a modified compound variable that 1) combines ethnicity and migrant status, and 2) Groups all individuals who had reported ethnicity as “White British” into a single reference group. Note that there were 204 individuals who reported being White British and not UK born, and a further 3,218 reporting to be White British with a missing migration status. These two groups were added to the existing 7,786 who reported being White British + UK Born and are listed below simply as “White British”.

|  | Experienced obstacles/barriers when accessing health services |  |  |
| --- | --- | --- | --- |
|  | Model adjusted for age, sex and ethnicity | Model adjusted for age, sex and migration status | Model adjusted for age, sex and ethnicity-migration compound variable |
| **Ethnicity** |  |  |  |
| White British | Ref | x | x |
| Minority ethnic | 1.1 (1.0-1.3) | x | x |
| **Place of birth** |  |  |  |
| UK Born | x | Ref | x |
| Not UK born | x | 1.0 (0.9 -1.2) | x |
| **Ethnicity + migration status\*\*** |  |  |  |
| White British | x | x | Ref |
| Minority ethnic-Missing migration status | x | x | 1.0 (0.8 -1.3) |
| Minority ethnic - Not UK Born | x | x | 1.03 (0.9-1.2) |
| Minority ethnic - UK Born | x | x | 1.4 (1.1-1.9)* |

\*p-value ≤ 0.05 or less.
\*\*There were 11 participants for whom ethnicity was missing, 9 of which reported being UK born and 2 of which were not UK born. While these were retained in the model, they are not presented here due to their small numbers. There were 35 participants who were excluded from the model for whom both ethnicity and migration status were missing.

### Reasons for difficulty accessing healthcare

Most participants (97.9%, 4,474/4,571) who reported difficulty accessing healthcare specified one or more reasons for this difficulty. The most commonly reported barrier was “Services disrupted or cancelled due to the COVID-19 pandemic” (72.0%, 3,291/4,571). Figure 1 shows the top ten reported barriers by IMD quintile, ethnicity and migrant status and excluding this first most common one so as to better illustrate differences in the remaining barriers. Appendix 1 shows the distribution of reasons by IMD quintile, ethnicity and migration status whereas appendix 2 shows the frequency of each reason across all groups). The second and third most frequently described reasons were problems with digital and/or telephone access (21.9%, 999/4,571), and lack of flexibility of appointments (21.7%, 990/4,571).

**Figure 1:**
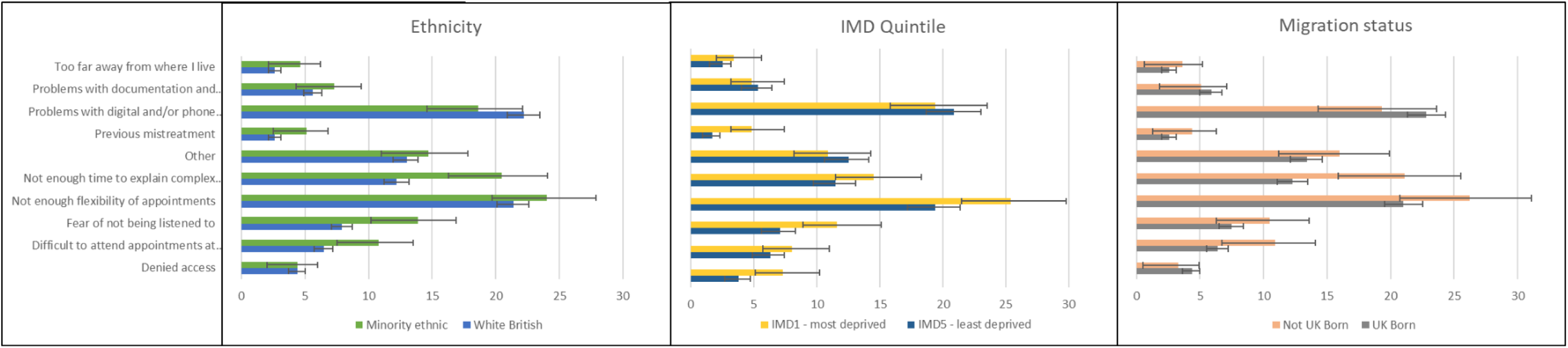
This bar chart shows the percentage of participants in each group who selected each barrier to healthcare access. This figure purposely excludes the most commonly selected reason, “Services having been disrupted or cancelled due to the COVID-19 pandemic” so as to better illustrate the differences between the remaining, top ten selected barriers.

Regarding the less frequently described reasons, participants from a minority ethnic background were more likely than White British participants to report difficulty attending appointments at the times they were offered (10.7% [95% CI 8.1 to 14.1] vs. 6.5% [5.8 to 7.3]), fear of not being listened to (13.9% [10.9 to 17.6] vs. 7.9% [7.1 to 8.7]), not having enough time to explain complex needs (13.9% [10.9 to 17.6] vs. 7.9% [7.1 to 8.7]), previous mistreatment (5.1% [3.4 to 7.7] vs. 2.6% [2.1 to 3.1]), and previous experience of discrimination or stigma (3.4% [2.0 to 5.7] vs. 1.5% [1.2 to 2.0]). Migrants were more likely than non-migrants to cite not having enough time to explain complex needs (21.1% [16.7 to 26.3] vs. 12.2% [11.1 to 13.5]). There was also an upwards trend with increasing socioeconomic deprivation for likelihood to cite fear of not being listened to (11.6% [8.9 to 15.1] in the 1st IMD quintile vs. 7.1% [5.9 to 8.6] in the 5th), previous mistreatment (4.8% [3.2 to 7.4] in the 1st quintile vs. 1.7% [1.1 to 2.5] in the 5th) and being denied access (7.3% [5.1 to 10.2] in the 1st quintile vs. 3.8% [2.9 to 4.9] in the 5th).

## Discussion

### Main findings

We examined the characteristics associated with reporting difficulties in accessing healthcare during the year 2021 in England and Wales, and the obstacles/barriers related to this. Most (78.6%) of the participants who completed the Access survey reported trying to access healthcare, of which over a third (37.4%) reported facing at least one barrier in doing so. We found higher proportions of this outcome among minority ethnic participants and those residing in more deprived IMD quintiles, while migrants and non migrants reported this outcome at broadly similar levels, although evidence of the association between ethnicity/migration and barriers to accessing healthcare after adjusting for age and sex weakened. Our model, which included a modified compound variable for ethnicity plus migrant status, showed a stronger association with the primary outcome for those who were both minority ethnic and UK born than other permutations within the same variable. The most frequently reported barrier across groups was cancellation or disruption in healthcare services during the COVID-19 pandemic. We also found that participants from minority ethnic backgrounds reported difficulty accessing healthcare - particularly attending appointments at the times offered and insufficient flexibility of appointments - more than their White British counterparts. This could be for a range of reasons including household make up and occupation types. For example, Census data shows that higher proportions of ethnic minority adults live with dependent children, compared with White British people^15^, likely leading to greater childcare responsibilities.

### What is already known on this topic

Other studies have suggested that minoritised groups faced different - and sometimes more - barriers to healthcare than their counterparts^10–13^. A previous study using administrative data found that migrants were less likely to use primary care than non-migrants before the pandemic and the first year of the pandemic exacerbated this difference^11^. In contrast, our survey found that broadly similar proportions of migrants and non-migrants (39.3% [35.8 - 43.0] vs. 36.8% [35.7 - 37.8], respectively) self-reported difficulty accessing healthcare. This may suggest a mismatch between these groups of perceived vs actual need, use and access to healthcare, and warrants further research.

Maddock et al reported evidence from 12 UK population-based longitudinal studies wherein both ethnic minorities and those in lower occupational classes were more likely to report health disruptions during the pandemic^13^. Upon stratification by shielding status, the strength of association with any healthcare disruptions increased for ethnic minority groups. Our study findings also support this, however in examining the domains of disruptions (medications, appointments or procedure), these associations became less clear and the actual reasons for healthcare disruption from the view of the patient was out of scope.

Pre-pandemic research has suggested many potential barriers to healthcare access for minority groups including language, cultural differences, discrimination and experiences of structural racism, the latter two of which are supported by our study ^16,17^ . However, these and the other barriers reported by participants in this study exist to some extent on all levels, and the evidence suggests disadvantages in healthcare access are compounded at the intersection of ethnicity, migration and socio-economic status^18^.

### What this study adds

A previous study examining hospital admissions during the pandemic according to area deprivation and proportion of ethnic minorities suggested that falls in admissions found were “consistent with evidence from the UK and the USA that ethnic minorities have been more likely to avoid seeking care during the pandemic”^12^ . However, this may be at least partly explained by finding in our study, where some of the barriers/obstacles reported by ethnic minority and migrant participants could collectively be considered as previous negative experiences with either the health service, or with institutions in general. For instance, people from ethnic minority backgrounds were more likely than their White British counterparts to select “fear of not being listened to”, “not having enough time to explain complex needs”, “previous mistreatment”, and “previous experiences of discrimination or stigma” as barriers. A similar pattern was found for migrants compared to UK born participants; however, the latter should be interpreted with caution, as it includes small numbers (appendix 1). Nonetheless, this points to a need to ensure that people from ethnic minority backgrounds can present to health services without these fears and concerns, especially because accessing preventative and timely care before it cascades into bigger health problems is important for both patients, and the cost efficiency of health services.

An apparent gradient was also found in IMD quintiles with individuals living in more deprived areas reporting difficulty accessing healthcare more frequently than those living in less deprived areas - a finding that is supported by previous research^19^. This should be considered alongside the findings for minority ethnic groups, especially given recent figures showing that ethnic minority groups are more likely than White British people to live in the most overall deprived 10% of neighbourhoods in England (with the exception of individuals who are of Indian, Chinese, White Irish and White Other ethnicity)^20^.

### Limitations

The Virus Watch study is a large and broadly diverse cohort in terms of demographic characteristics such as age, sex, ethnicity, and socioeconomic status, as well as being well-distributed across the geographic areas of England and Wales.

However, participants responding to the January 2022 Access survey were more likely to be over 65, to be of White British ethnicity, to be female, and to be of higher socioeconomic status than the general population in England and Wales. This raises concerns that the sample has higher representation of non-minoritised individuals, and therefore higher likelihood of multiple advantage than disadvantage, when it is the latter that is of interest to our research questions. Nonetheless, the use of confidence intervals in our descriptive analysis and multivariable regression should serve to mitigate this issue as these methods account for differing sample sizes and show, with reasonable reliability, the direction of association.

We used a binary variable for minority ethnicity status in order to enable sufficient sample size for comparisons, and as a result we have been unable to examine more detailed comparisons between specific minority ethnic groups. Although this aggregation was acceptable (although not preferred) to the Virus Watch advisory group for pragmatic reasons, it could mean that important nuances in barriers to healthcare were not described and further work with greater sample sizes should seek to explain these.

Other than the issues identified for participants from minority ethnic backgrounds, significant evidence of disproportionate barriers specific to migrants were not found in the present analysis, likely due to the Virus Watch recruitment process and study population. The requirement for the lead householder of a participating household to have Internet access and understand English well enough to fill in the surveys may have meant that difficulties with digital access and language issues were likely to be underestimated for migrants in the UK. In other words, while the shift to remote and digital access during the COVID-19 pandemic is likely to have exacerbated these difficulties for these groups, this is probably underestimated in this study due as participants were pre-selected as having digital access.

This study found that most individuals attributed barriers to pandemic-related disruptions.. The NHS constitution states that its services are available to all, regardless of personal characteristics, with access based on clinical need. However, this study found that during the COVID-19 pandemic, participants from ethnic minority backgrounds and those living the most deprived areas experienced barriers to accessing healthcare to a greater extent than their White British counterparts and those living in less deprived areas, respectively. As services continue to manage ever present backlogs of care, it is essential these inequities in access are examined comprehensively together, and that the barriers are addressed.

## APPENDICES

## Appendix 1: Breakdown by groups of reasons given for difficulty accessing healthcare

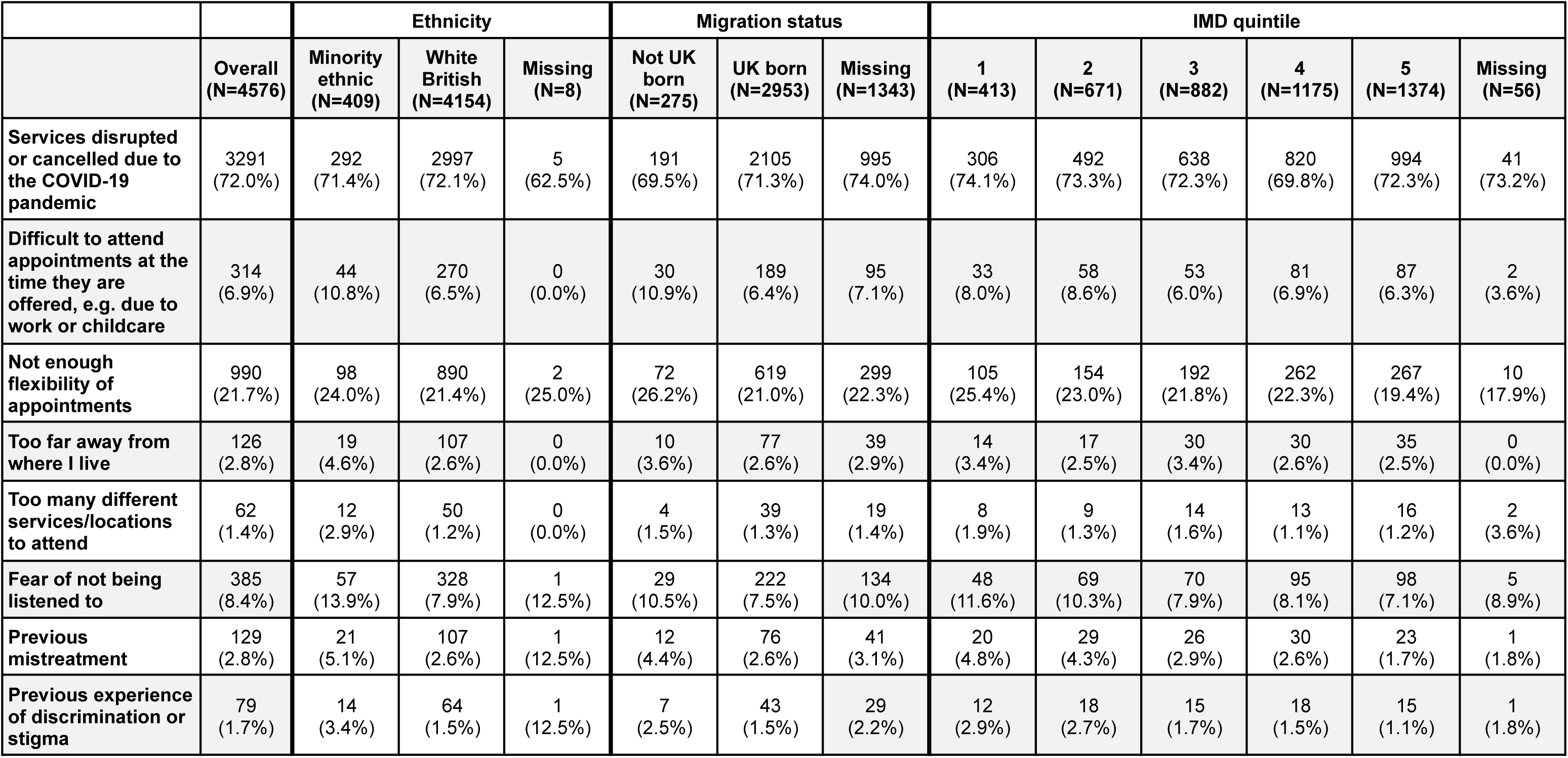
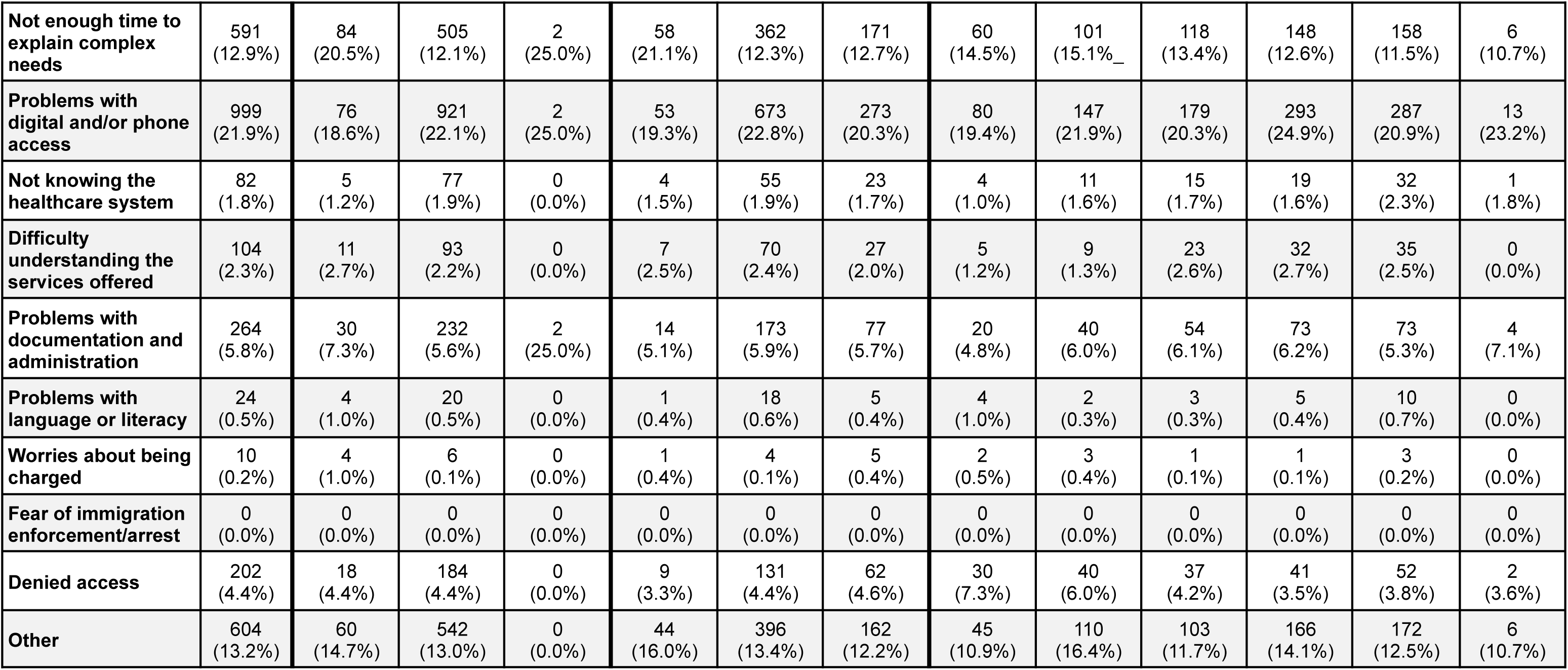

## Appendix 2: Figure: All reasons selected for difficulty accessing healthcare

**Figure:**
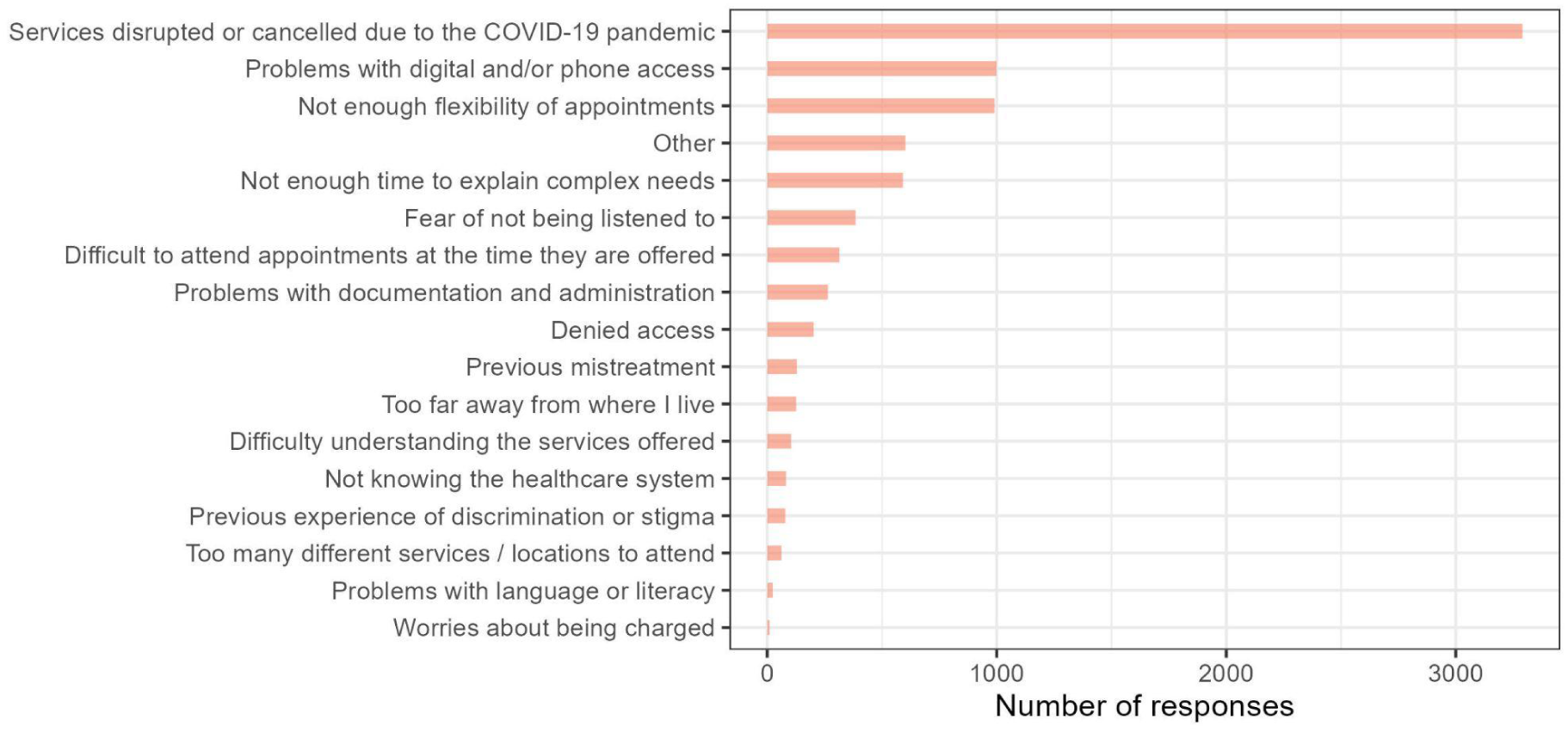
All reasons selected for difficulty accessing healthcare

## Appendix 3: Models age, sex, ethnicity and migration

|  | Experienced obstacles/barriers when accessing health services |  |  |  |
| --- | --- | --- | --- | --- |
|  | Model adjusted for age, sex and ethnicity | p-value | Model adjusted for age, sex and migration status | p-value |
| **Sex** |  |  |  |  |
| Male | - |  | - |  |
| Female | 1.4 (1.3-1.5) | <0.001 | 1.3 (1.1-1.4) | <0.001 |
| **Age Group** |  |  |  |  |
| 16-24 | - |  | - |  |
| 25-44 | 1.7 (1.2-2.5) | 0.005 | 2.03 (1.3-3.4) | 0.005 |
| 45-64 | 1.4 (1-2.1) | 0.029 | 1.5 (1-2.5) | 0.068 |
| 65+ | 1.1 (0.8-1.6) | 0.5 | 1.1 (0.7-1.8) | 0.6 |
| **Ethnicity** |  |  |  |  |
| White British | - |  | x |  |
| Minority ethnic | 1.1 (1.0-1.3) | >0.9 | x |  |
| **Place of birth** |  |  |  |  |
| UK Born | x |  | - |  |
| Not UK born | x |  | 1.0 (0.9 -1.2) | 0.7 |

## Appendix 4 Models with age, sex, deprivation and the “ethnicity-migration” compound variable

Appendix 4 - Models with age, sex, deprivation and the “ethnicity-migration” compound variable
|  | Fully adjusted ORs (95% CIs) | p-value | Age, sex & ethnicity x migration interaction variable | p-value | N |
| --- | --- | --- | --- | --- | --- |
| **Sex** |  |  |  |  |  |
| Reference | - |  | - |  | 5,119 |
| Female | 1.4 (1.3-1.5) | <0.001 | 1.4 (1.3-1.5) | <0.001 | 7,072 |
| **Age Group** |  |  |  |  |  |
| 16-24 | - |  | - |  | 142 |
| 25-44 | 1.7 (1.2-2.6) | 0.005 | 1.7 (1.2-2.6) | 0.005 | 730 |
| 45-64 | 1.5 (1-2.2) | 0.029 | 1.5 (1-2.1) | 0.042 | 4,688 |
| 65+ | 1.1 (0.8-1.7) | 0.5 | 1.1 (0.8-1.6) | 0.6 | 6,631 |
| **IMD Quintile** |  |  |  |  |  |
| 5 (least deprived) | - | - | x | x | 3,856 |
| 4 | 1.1 (1-1.2) | 0.3 | x | x | 3,165 |
| 3 | 1.0 (0.9-1.1) | >.9 | x | x | 2,420 |
| 2 | 1.1 (1-1.3) | 0.075 | x | x | 1,699 |
| 1 (most deprived) | 1.4 (1.2-1.6) | <0.001 | x | x | 904 |
| **Ethnicity** |  |  |  |  |  |
| White British | - |  |  |  | 11,208 |
| Minority ethnic-Missing migration status | 1 (0.8-1.3) | >0.9 | 1.0 (0.8 -1.3) | 0.9 | 257 |
| Minority ethnic - Not UK Born | 1 (0.8-1.2) | >0.9 | 1.03 (0.9-1.2) | 0.8 | 493 |
| Minority ethnic - UK Born | 1.5 (1.1-1.9) | 0.006 | 1.4 (1.1-1.9) | 0.006 | 233 |

## Competing Interest Statement

AH serves on the UK New and Emerging Respiratory Virus Threats Advisory Group. All other authors declare no competing interests.

## Data Availability

We aim to share aggregate data from this project on our website and via a "Findings so far" section on our website: https://ucl-virus-watch.net/. We also share some individual record level data on the Office of National Statistics Secure Research Service. In sharing the data we will work within the principles set out in the UKRI Guidance on best practice in the management of research data. Access to use of the data whilst research is being conducted will be managed by the Chief Investigators (AH and RA) in accordance with the principles set out in the UKRI guidance on best practice in the management of research data. We will put analysis code on publicly available repositories to enable their reuse. We aim to share aggregate data from this project on our website and via a "Findings so far" section on our website: https://ucl-virus-watch.net/. We also share some individual record level data on the Office of National Statistics Secure Research Service. In sharing the data we will work within the principles set out in the UKRI Guidance on best practice in the management of research data. Access to use of the data whilst research is being conducted will be managed by the Chief Investigators (AH and RA) in accordance with the principles set out in the UKRI guidance on best practice in the management of research data.

## Notes

### Funding Statement

Virus Watch was supported by the Medical Research Council [Grant Ref: MC_PC 19070 and MR/V028375/1]. The study also received $15000 of advertising credit from Facebook to support a pilot social media recruitment campaign on 18 August 2020. The antibody testing was also supported by funding from the Department of Health and Social Care from February 2021 to March 2022. This study was also supported by the Wellcome Trust through a Wellcome Clinical Research Career Development Fellowship to R.W.A. [206602].

## References

1. ONS. Disparities in the risk and outcomes of COVID. Preprint at https://assets.publishing.service.gov.uk/government/uploads/system/uploads/attachment_data/file/908434/Disparities_in_the_risk_and_outcomes_of_COVID_August_2020_update.pdf.

2. Holman, D. et al. Can intersectionality help with understanding and tackling health inequalities? Perspectives of professional stakeholders. Health Res. Policy Syst. 19, 97 (2021).

3. Beale, S. et al. Occupation, Worker Vulnerability, and COVID-19 Vaccination Uptake: Analysis of the Virus Watch prospective cohort study. Vaccine 40, 7646–7652 (2022).

4. Beale, S. et al. Occupation, Work-Related Contact, and SARS-CoV-2 Anti-Nucleocapsid Serological Status: Findings from the Virus Watch prospective cohort study. bioRxiv (2021) doi:10.1101/2021.05.13.21257161.

5. Aldridge, R. W. et al. Household overcrowding and risk of SARS-CoV-2: analysis of the Virus Watch prospective community cohort study in England and Wales. Wellcome Open Res. 6, 347 (2021).

6. Stagg, H. R., Jones, J., Bickler, G. & Abubakar, I. Poor uptake of primary healthcare registration among recent entrants to the UK: a retrospective cohort study. BMJ Open 2, (2012).

7. Hamada, R. et al. Most GP surgeries refuse to register undocumented migrants despite NHS policy. The Bureau of Investigative Journalism (en-GB) https://www.thebureauinvestigates.com/stories/2021-07-15/most-gp-surgeries-refuse-to-register-undocumented-migrants.

8. Saunders, C. L. et al. Healthcare utilization among migrants to the UK: cross-sectional analysis of two national surveys. J. Health Serv. Res. Policy 26, 54–61 (2021).

9. Nazroo, J. Y., Falaschetti, E., Pierce, M. & Primatesta, P. Ethnic inequalities in access to and outcomes of healthcare: analysis of the Health Survey for England. J. Epidemiol. Community Health 63, 1022–1027 (2009).

10. Poorest get worse quality of NHS care in England, new research finds. Nuffield Trust https://www.nuffieldtrust.org.uk/news-item/poorest-get-worse-quality-of-nhs-care-in-england-new-research-finds.

11. Zhang, C. X. et al. Migrants’ primary care utilisation before and during the COVID-19 pandemic in England: An interrupted time series analysis. The Lancet Regional Health - Europe 20, 100455 (2022).

12. Warner, M. et al. Socioeconomic deprivation and ethnicity inequalities in disruption to NHS hospital admissions during the COVID-19 pandemic: a national observational study. BMJ Qual. Saf. 31, 590–598 (2022).

13. Maddock, J. et al. Inequalities in healthcare disruptions during the COVID-19 pandemic: evidence from 12 UK population-based longitudinal studies. BMJ Open 12, e064981 (2022).

14. Byrne, T., et al. Cohort profile: Virus Watch: Understanding community incidence, symptom profiles, and transmission of COVID-19 in relation to population movement and behaviour. medRxiv (2023) doi:10.1101/2023.01.31.23285232.

15. Families and households. https://www.ethnicity-facts-figures.service.gov.uk/uk-population-by-ethnicity/demographics/families-and-households/latest (2019).

16. Gazard, B. et al. Barrier or stressor? The role of discrimination experiences in health service use. BMC Public Health 18, 1354 (2018).

17. Hatzenbuehler, M. L., Phelan, J. C. & Link, B. G. Stigma as a fundamental cause of population health inequalities. Am. J. Public Health 103, 813–821 (2013).

18. Abubakar, I. et al. The UCL-Lancet Commission on Migration and Health: the health of a world on the move. Lancet 392, 2606–2654 (2018).

19. Moscelli, G., Siciliani, L., Gutacker, N. & Cookson, R. Socioeconomic inequality of access to healthcare: Does choice explain the gradient? J. Health Econ. 57, 290–314 (2018).

20. People living in deprived neighbourhoods. https://www.ethnicity-facts-figures.service.gov.uk/uk-population-by-ethnicity/demographics/people-living-in-deprived-neighbourhoods/latest (2020).

